# Monitoring of Schools and Colleges of Pharmacy Based on NAPLEX Passage Rate

**DOI:** 10.1101/2023.05.02.23289407

**Authors:** Natalia Shcherbakova, John M. Pezzuto

## Abstract

**Introduction:** We have evaluated two approaches of monitoring schools and colleges of pharmacy based on North American Pharmacist Licensure Examination® (NAPLEX®) passage rates. Historically, the Accreditation Council for Pharmacy Education (ACPE) has cited programs based on achieving a passage rate ≥2 SD standard deviations (SD) below the national average. Since the National Association of Boards of Pharmacy (NABP) no longer reports scores, this procedure is currently being reconsidered. Our supposition is that the failure rate of ≥2 SD below the average passage rate should be retained, but now, based on the national average passage rate. However, we further suggest this is not adequate, due to major variations in class size.

**Commentary:** We suggest the establishment of a ‘maximum acceptable failure count’, likely in the range of 20-25 failing graduates per class. Analyses of data from 2017-2019 indicate that this approach would lead to monitoring approximately 15% of existing programs that graduate approximately 40% of individuals failing NAPLEX, versus monitoring approximately 5% of programs that graduate approximately 9% of individuals failing NAPLEX.

**Implications:** The historical method of monitoring pharmacy programs with NAPLEX passage based on the national average ≥2 SD is not adequate, primarily due large variations in class size. In that accreditation standards are currently being revised (“Standards 2025”), this would be an ideal time to reconsider methods for selecting programs that warrant monitoring based on inadequate NAPLEX passage rates. We suggest the concept of ‘maximum acceptable failure count’ should be taken into account when identifying programs to be cited.

## Introduction

The importance of meeting professional accreditation standards by US colleges and schools of pharmacy is self-evident. The standards, as administered by the Accreditation Council for Pharmacy Education (ACPE), are comprehensive and well justified. The ultimate goal of professional accreditation is clearly stated in the Mission Statement of the ACPE: “By providing accreditation services and professional development activities, ACPE assures the quality of pharmacy education and training and supports the advancement of the pharmacy profession.” ^1^

While there are many requisite outcomes that validate the competence of a new PharmD graduate, passage rate on the NAPLEX is certainly one that may be viewed as a ‘gold standard.’ As such, it is clear that the passage rate of graduates is one important parameter that is indicative of the success of a pharmacy program. It may be suggested that pharmacy programs should be evaluated by different approaches in relation to NAPLEX pass rates, i.e., monitoring of first time test takers who are successful in passing the NAPLEX vs. monitoring of ultimate passage rate of test takers following two or more attempts. In our view, monitoring of first time test takers is of greatest relevance, since this should be most highly indicative of the preparedness of a graduate provided directly by the respective college or school of pharmacy.

It is likely that every school or college of pharmacy aspires for their graduates to achieve a 100% pass rate when taking the NAPLEX. Since attainment of the goal is rarely observed, especially by first-time test takers, in essence, the question becomes what is an acceptable rate of failure. Historically, ACPE has cited programs when the percentage of graduates failed NAPLEX at a rate of two standards deviations below the national average. The rationale of this approach is stated as follows: “ACPE uses two standard deviations as a cut-off value as this provides assurance that the monitoring is addressing lower performing programs given that 95% of data values lie within two standard deviations. This also adjusts for year-to-year variation in NAPLEX scaling scores. Review of this data and the program’s response can lead to additional monitoring or a finding of partial or non-compliance with an accreditation standard”.^2^

More recently, when NABP changed reporting from a numerical score to pass/fail result (January 2021), the ACPE announced mean scaled scores could no longer be monitored. Although the percentage pass rate will continue to be monitored, the citation paradigm of -2 SD would no longer be applied.^3^ Relevant to this point, pharmacy programs are currently judged by the guiding principles of “Standards 2016”, but revision is underway with the release of new standards being planned for June 2024 and taking effect on July 1, 2025 (“Standards 2025”). This presents an obvious opportunity to revisit monitoring of NAPLEX passage rates. Indeed, in an update provided by ACPE in February 2022^4^, it was stated “The Board is considering methods to make reporting more accurate. They are also discussing whether there should be a required permanent benchmark pass rate or bright line for NAPLEX versus using the current measure of 2 standard deviations below the average. ACPE staff will be reaching out to the academy for feedback on these issues.”

## Commentary

Based on these circumstances, we thought it would be of interest to explore various pathways going forward in regard to monitoring pharmacy programs based on NAPLEX passage rate. One straightforward idea would be to simply consider the average passage rate, set this as a bar, and monitor programs falling at or lower than the average. Recently, Nau et al.^5^ have evaluated the rankings of pharmacy programs, with some emphasis on NAPLEX scores being a key determinant, and cited a survey^6^ in which an 80% pass rate was proposed as the minimum threshold that all schools should achieve. However, arbitrary selection of a fixed pass rate percentage presents a disproportional discrepancy. For example, consider a hypothetical situation where the average pass rate were 80%, and 70% were set as the monitoring point. In this case, Program A, having a class size of 40 with a 70% pass rate would be monitored (12 failures), whereas Program B, with a class size of 200 and a pass rate of 80% would not be monitored (40 failures). The number of failures emanating from Program B would actually be equal to the entire class size of Program A. In this context, the monitoring of Program A and not Program B appears incongruent. Accordingly, consideration should be given to methods other than an arbitrary minimally acceptable percent pass as a potential criterion for monitoring.

A primary reason for conducting the current analysis was the change in NABP reporting from a score to pass/fail, which naturally leads us to using percentage pass rate and total number of test takers. With these data in hand, it is not a complicated matter to follow past practice of determining what programs would warrant monitoring using the historical process of the ACPE (mean pass rate – 2 SD) as the key determinant. We started this analysis with data available from 2018.^7^ Following the historical monitoring paradigm, out of 134 schools with first time pass rates, the mean pass rate was 89.125%. Minus two standard deviations yields a value of 73.12%, which should lead to monitoring of seven out of the 134 programs (5.2%) (Table 1). It seems reasonable that this practice be retained, i.e., programs with pass rates at or below the average national pass rate minus 2 SD should be monitored, irrespective of the size of the program.

**Table 1:**
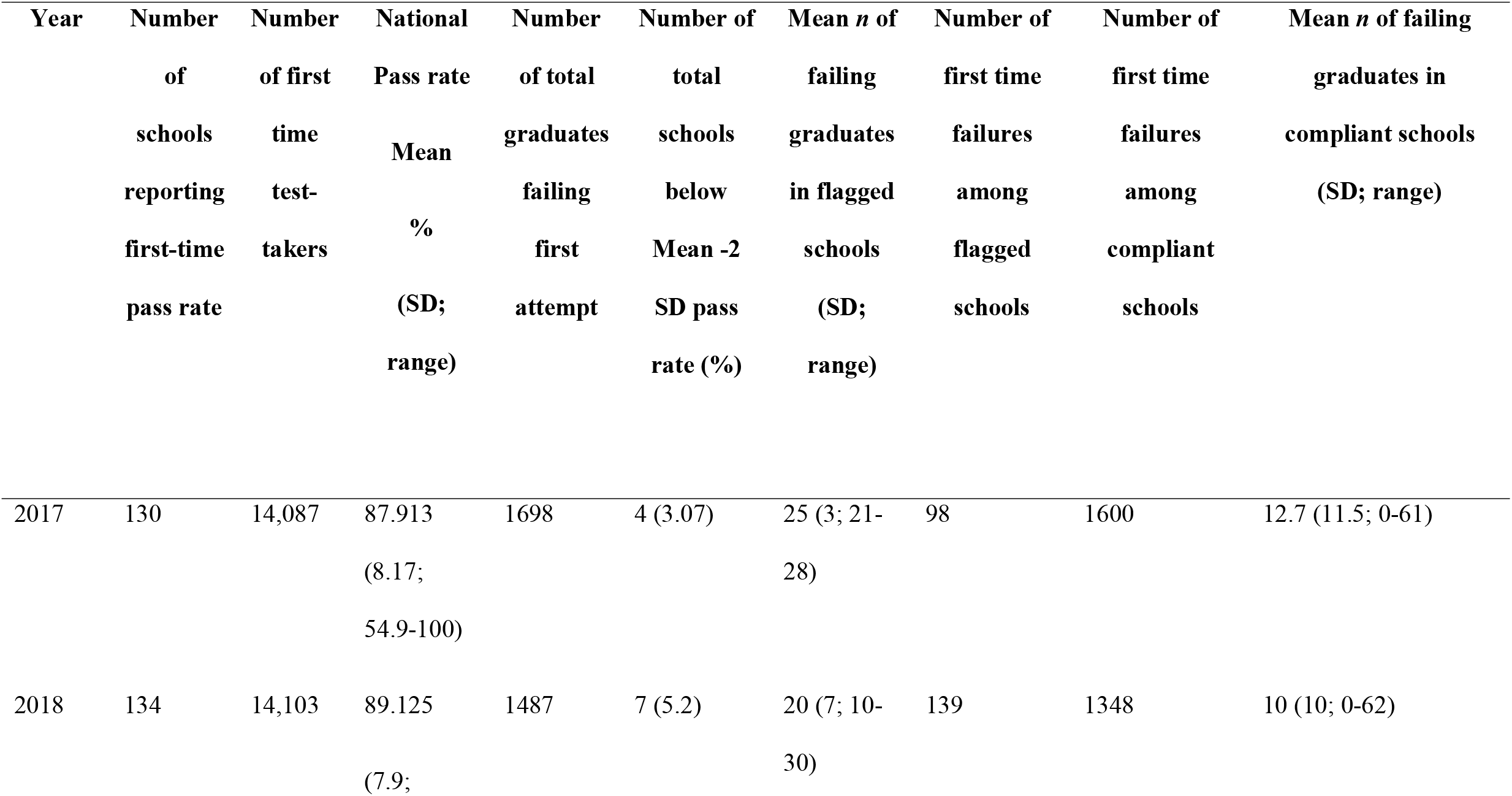
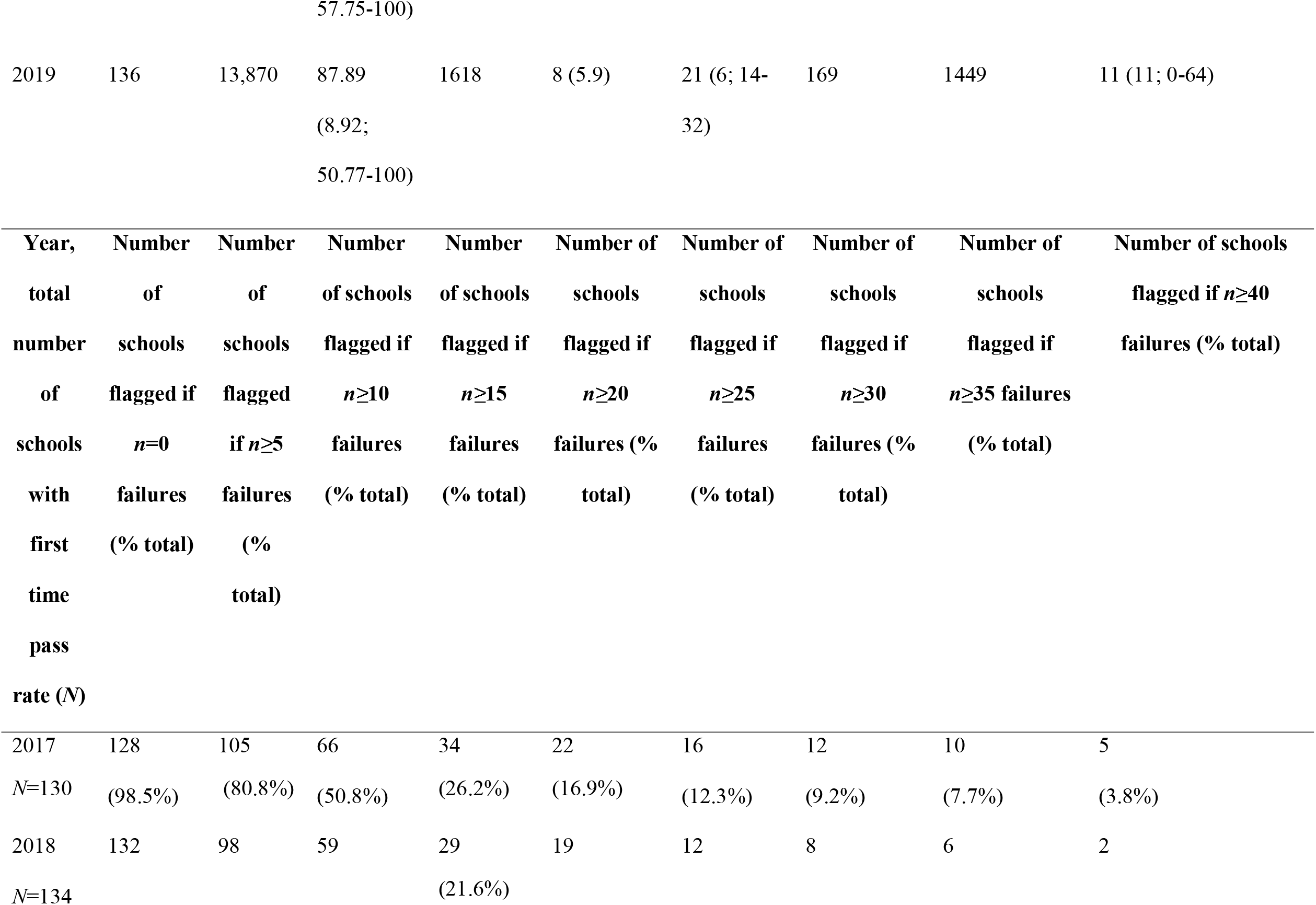
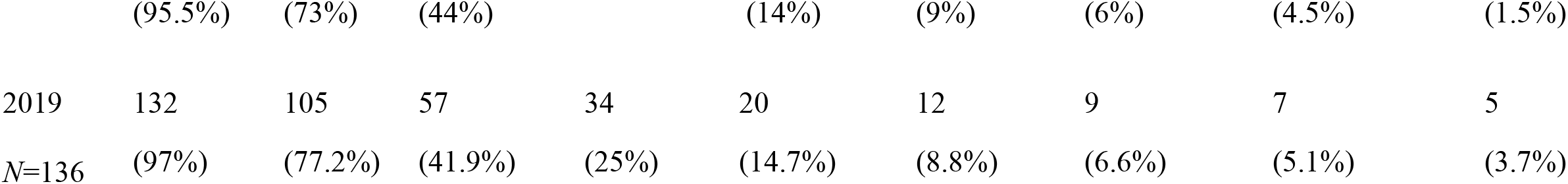
National North American Pharmacist Licensure Examination statistics and average number of graduates failing in flagged vs. not flagged schools according to Accreditation Council for Pharmacy Education rule of mean minus 2 SD and number of schools being flagged based on potential acceptable count for number of first-time failures.

However, in absolute terms, the total number of individuals failing NAPLEX who graduated from the seven programs that would be subjected to monitoring was 139. Of the remaining programs, not subjected to monitoring, the total number of failures was 1348 (9.7-fold greater) (Table 1). Our original intent in performing this exercise was to explore application of the historical monitoring paradigm to the current situation in which NABP provides only pass/fail data. However, the fact of 139 failures leading to program monitoring, whereas 1348 failures would not be explicitly subjected to monitoring, gave us pause. So next, we thought it would be of interest to examine alternative theoretical approaches that may more fully capture the impact of passage rate on program monitoring, to serve as an augment to the practice of only monitoring programs falling at least 2 SD below the average.

Again focusing on 2018, the mean number of graduates failing from the seven programs with pass rate below two SD of the mean was 20, compared to a mean of 10 graduates in all of the non-monitored programs (Table 1). We decided to test what proportion of schools would need to be monitored if the threshold were based on a count of the actual failures rather than a pre-specified acceptable pass rate. We applied a simple approach based on the absolute count of first time failures. As a straightforward example, with a program of *n* first-time takers, where *n* = 100, with a pass rate of 70%, we know that 30 graduates failed [*n* x (1-pass rate)]. Next, we considered what may be termed “maximal acceptable failure count.” Considering the data from 2018, setting the maximum allowable failure rate at 0, 5 or 10 yields a monitoring rate of between 95-44% of programs (Table 1). Setting the maximum acceptable failure count at 30, 35 or 40 yields a monitoring rate between 6-1.5%. Common sense tells us that neither of these boundaries is reasonable. So, the question becomes, what value of allowable failures found between these boundaries would be fair to graduates, fair to programs, and meet the intrinsic objectives of the accrediting body to use NAPLEX pass rate as a quality indicator.

First, we may consider comparing the approach of using the average pass rate -2 SD paradigm with that of allowable count of failures. Again, evaluating the 2018 data^7^, allowable failures would need to approach 20 in order to capture four of the seven of the programs identified for monitoring, mainly due to smaller class sizes, albeit with high failure rates, using pass rate - 2 SD paradigm (Table 2). Hence, if we apply 20 failing graduates as the maximum allowable failure count for all programs, then 19 programs (14%) would be flagged for monitoring, which includes four of the seven programs originally identified due to passage rates below 2 SD of the national mean (Table 2). Thus, a total of 21 programs would be subject to monitoring, including all programs with pass rate below 2 SD of the national mean, and an additional 15 programs with 20 or more graduates failing the exam. In this case, at the 20 failures threshold, of the 1487 graduates failing NAPLEX, 606 graduates (40.8%) would be from programs that would be subjected to monitoring. Increasing the maximum acceptable failure count to 25 reduces the number of programs subject to monitoring to 12, which includes only two of the original seven selected on the basis on low passage rate. In this case, given that a grand total of 17 programs would be monitored, of the 1487 graduates failing NAPLEX, the number of graduates emanating from monitored programs would be reduced to 519 (34.9%). Further increases in the maximum allowable failure count to 30, 35 and 40, decreases the number of programs subject to monitoring to 15, 13 and 9, respectively, and decreases the proportion of failing graduates emanating from monitored programs to 463 (31.1%), 396 (26.6%) and 247 (16.6 %), respectively.

**Table 2:**
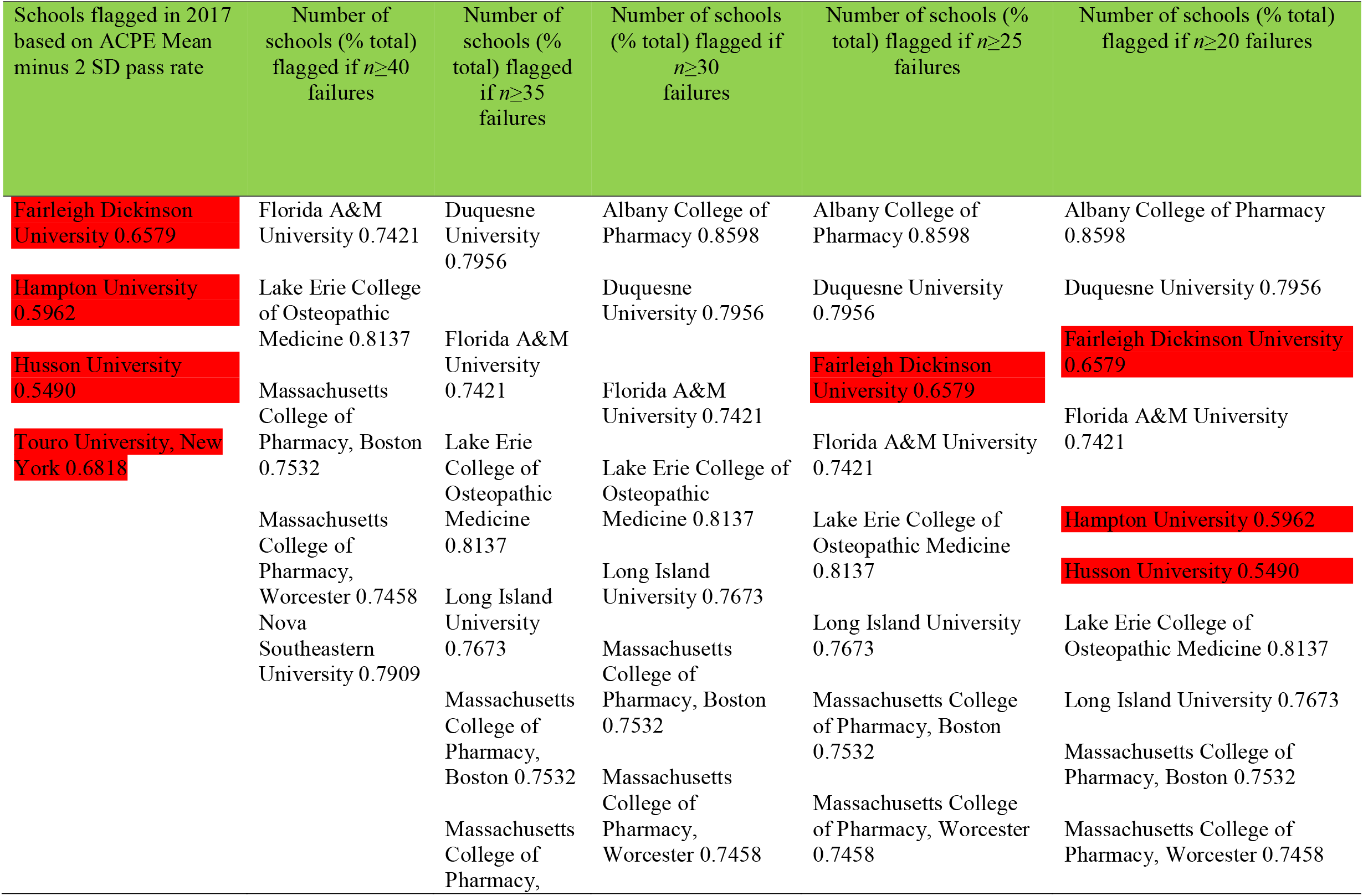
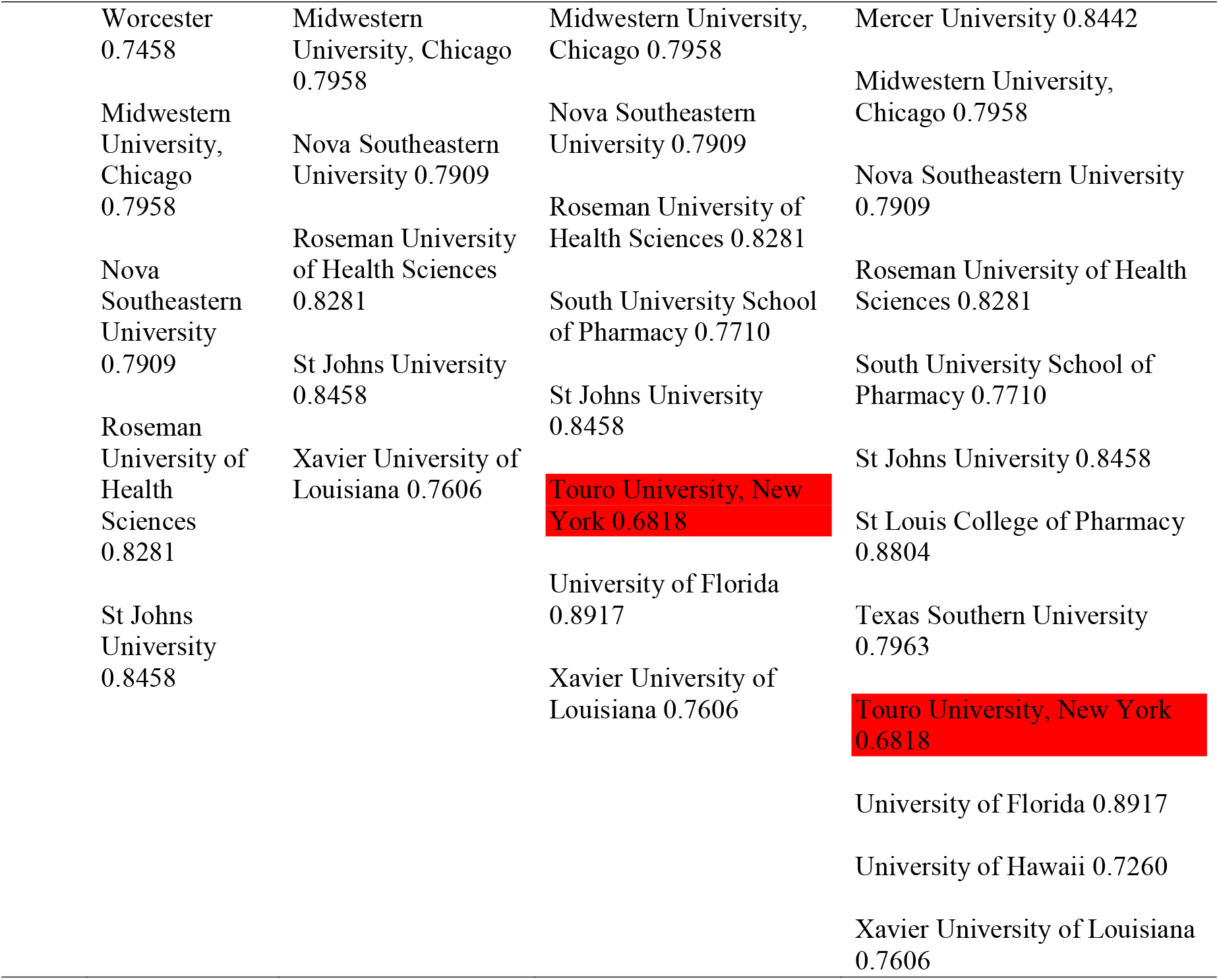
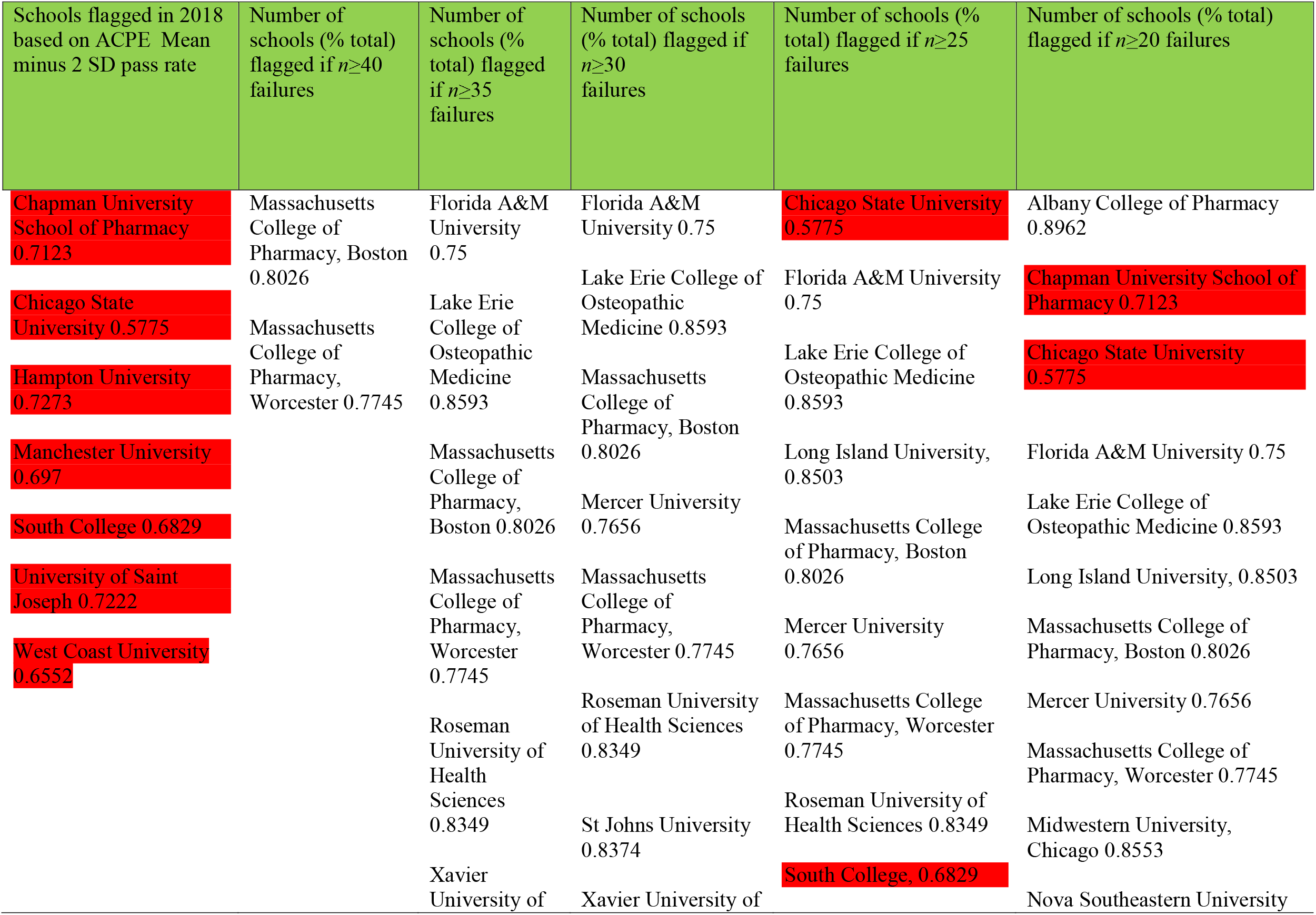
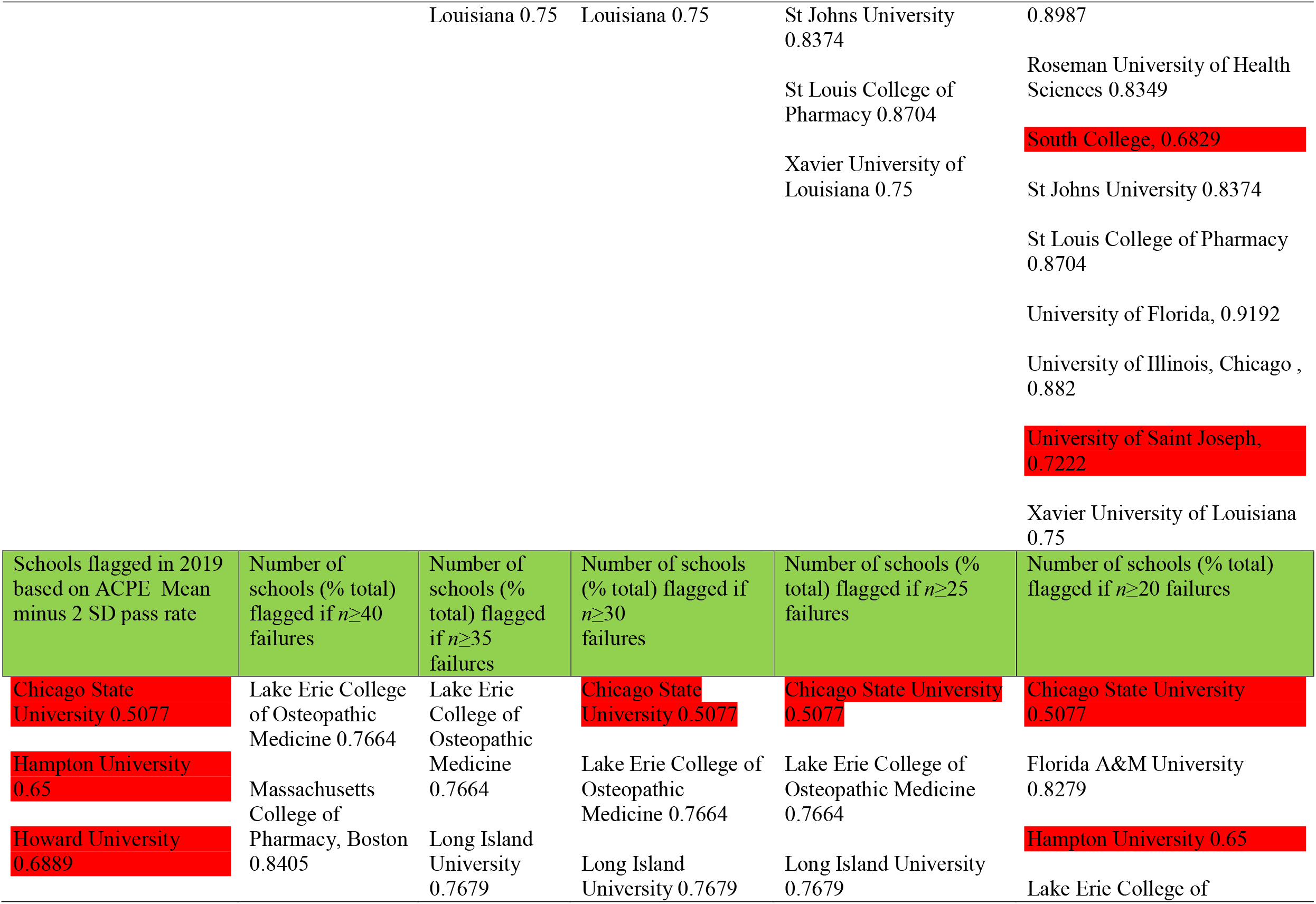
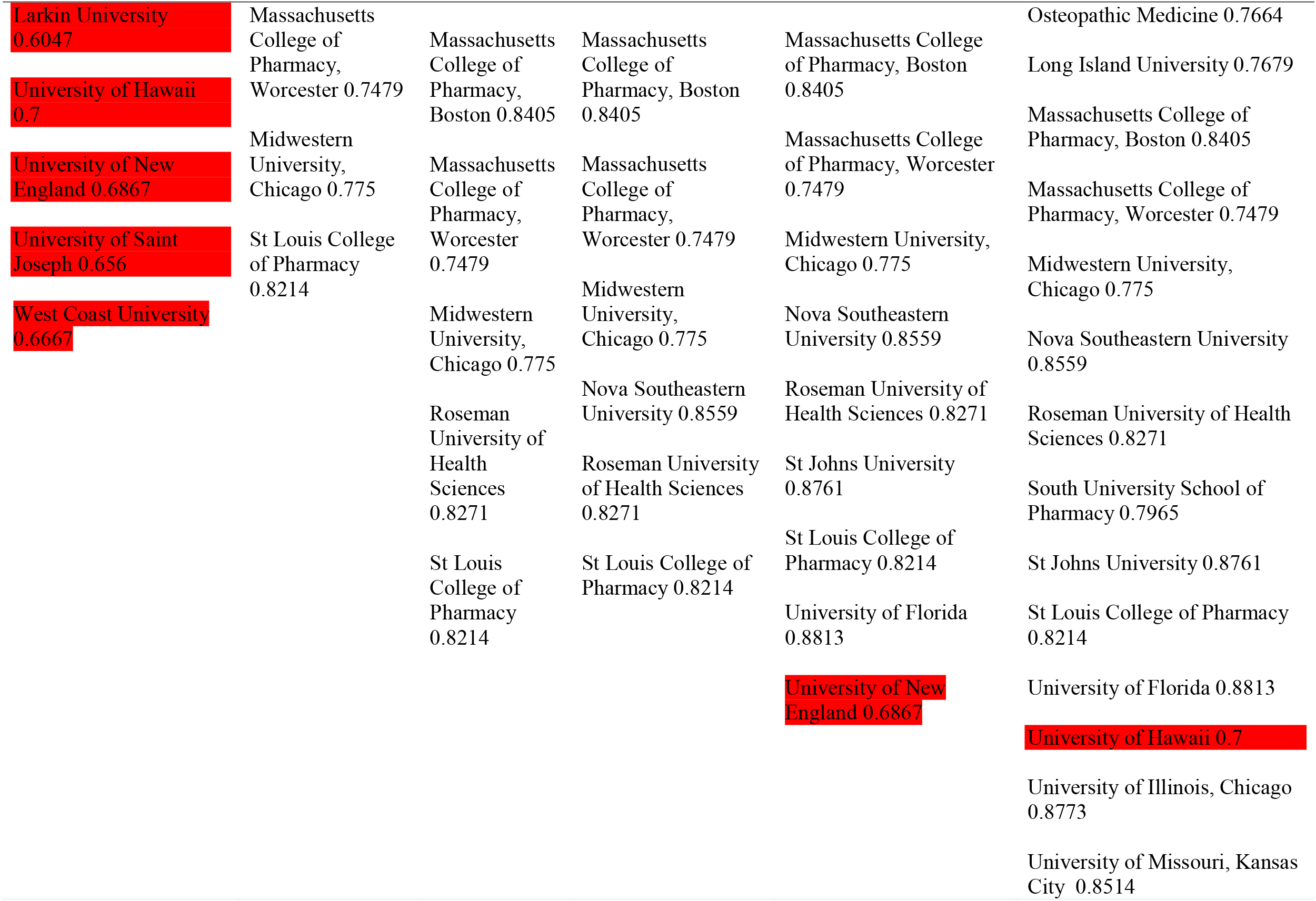
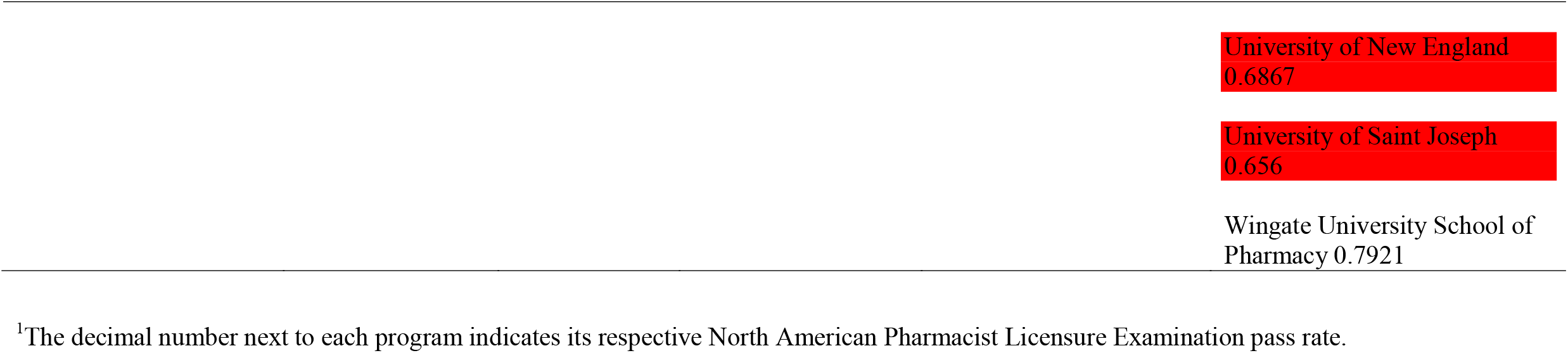
Comparison of ACPE school flagging rule with natural failure count approach.^1^

Analyses similar to those described above with 2018 data were performed with data from 2017 and 2019 and are summarized in Tables 1 and 2.^7^ In both cases, extremely similar trends were observed, with no major divergence in the results. Accordingly, this leads us to believe pass/fail data, without raw scores, are adequate but insufficient for justly identifying schools and colleges of pharmacy that warrant monitoring by the ACPE. Additional factors need to be taken into account, reflective of class sizes and the absolute number of failures.

## Implications

In sum, our supposition is that monitoring of pharmacy programs based on NAPLEX pass rate is a time-tested indicator of programmatic success that can be successfully supplemented with an additional approach using absolute counts of failures. Unacceptably low passage rate, relative to national norms, clearly must correlate with insufficiency in meeting one or more of the various comprehensive accreditation standards. Historically, programs were solely selected for monitoring based on a percentage pass rate, but we do not believe this adequately reflects the intended goal of the process. Simply put, the great majority of individuals failing NAPLEX are graduates of programs not historically subjected to monitoring. We present herein a supplemental to the pass rate approach that should promote greater equity and accountability. We note some programs that would be identified for monitoring using both the traditional and supplemental approaches described herein are programs that have relatively high passage rates yet add substantially to the pool of first-time failures. Hence, the absolute number of failures merits greater attention than heretofore brought to bear.

Finally, our intent in analyzing NAPLEX passage rate and the monitoring of pharmacy programs was not intended to be critical of past procedures, nor to be prescriptive for future procedures. Obviously, this is outside the realm of our authority. Our aspiration is to share factual and objective data that may be taken into account by pharmacy programs as part of self-assessment and, perhaps, have some influence on the ongoing process of formulating “Standards 2025.” It seems clear to us, however, that only utilizing a fixed pass rate to identify schools or colleges of pharmacy for monitoring is not adequate. As a corollary, nor is the approach of only using the absolute number of failures adequate, due to major variations in the size of graduating classes. At the same time, we do not advocate holding back graduates based on passing ‘high stakes’ examinations that would predict success on NAPLEX passage rates. This is akin to gaming the system which bears little resemblance to providing a high quality education in the profession of pharmacy.

## Data Availability

All data in the present study are in public domain.

https://nabp.pharmacy/wp-content/uploads/2019/03/NAPLEX-Pass-Rates-2019.pdf

## Contribution to literature

First time passage rate of Doctor of Pharmacy (PharmD) graduates on the North American Pharmacist Licensure Examination® (NAPLEX®) is key indicator of programmatic success. Historically, schools and colleges of pharmacy would be monitored by the Accreditation Council for Pharmacy Education (ACPE) if the passage rate of the program fell two standard deviations below the national average. As presented in this Commentary, it does not appear this approach fully captures the intent or spirit of the monitoring paradigm. We present an alternative approach that seems superior when used in conjunction with the current method. This analysis is especially timely since the professional standards adjudicated by the ACPE are currently under revision.

